# Bibliometric Analysis of Perioperative Organ Injury: Insights into the State of Basic Science Research

**DOI:** 10.1101/2024.09.19.24313978

**Authors:** Jinlian Wang, Hui Li, Changqing Ju, Yafen Liang, Holger Eltzschig, Hongfang Liu

## Abstract

Perioperative organ injury represents a significant clinical challenge, leading to severe and often long-term complications for patients undergoing surgery. Despite its critical importance, the basic science underlying these injuries remains underexplored. This study employs bibliometric analysis to assess the research landscape of perioperative organ injury, focusing on key conditions such as myocardial infarction, ischemia-reperfusion injury, heart failure, acute respiratory distress syndrome (ARDS), perioperative lung injury, liver ischemia-reperfusion injury, ischemic acute kidney injury, and perioperative stroke. By analyzing publication trends, citation patterns, and research themes, this study aims to provide insights into the current state of basic science research in this field, highlighting gaps and opportunities for future investigations.

## Introduction

Perioperative organ injury is a significant concern in surgical practice, often contributing to substantial morbidity and mortality. Common manifestations include neurological complications ^1^, myocardial ischemia ^2^, acute kidney injury ^3^, respiratory failure ^4^, intestinal dysfunction ^5^, and hepatic impairment ^6^. In Europe and the USA, postoperative mortality remains unexpectedly high, and if classified as its own category ^7,9^, perioperative mortality would rank as the third leading cause of death worldwide, after ischemic heart disease and cancer. More than 4 million patients die within 30 days of surgery annually, representing 7.7% of global deaths. Current treatment options are limited to supportive care, with no pharmacological therapy proven to prevent or reverse organ failure in clinical trials ^7^. Understanding the underlying mechanisms of organ dysfunction through basic science research is crucial to identifying new therapeutic targets and developing effective treatments ^10-13^.

Despite the significant impact of perioperative organ injury, there is a noticeable lack of basic science research focused on these mechanisms. A comprehensive understanding of these processes is essential to improving clinical practice and patient outcomes, but the vast scope of available literature makes it difficult to identify the most relevant research. A more systematic and focused approach is needed.

Given the complexity of perioperative organ injury, targeted literature searches using terms such as myocardial infarction, ischemia-reperfusion injury, heart failure, ARDS, perioperative lung injury, liver ischemia-reperfusion injury, ischemic acute kidney injury, and perioperative stroke are essential ^12-17^. Narrowing the focus to these specific conditions will enable a bibliometric analysis to highlight the current state of basic science research and identify areas requiring further investigation.

This study aims to perform a bibliometric analysis of the literature on perioperative organ injury, with an emphasis on the basic science aspects. The goal is to uncover key research trends, influential studies, and gaps in the field, providing a foundation for future research directions.

## Methods

### 1. Data collection

We searched PubMed and Web of Science (WOS) Core Collection database to collect publications related to 8 perioperative organ injury conditions: myocardial infarction, ischemia-reperfusion injury, heart failure, acute respiratory distress syndrome, perioperative lung injury, liver ischemia-reperfusion injury, ischemic acute kidney injury, and perioperative stroke at the day of Aug 28^th^ 2024. All searches and downloads were performed within one day to avert biases originating from daily database updating. The search strategies were presented, as shown in **Figure 1**.

**Figure 1.**
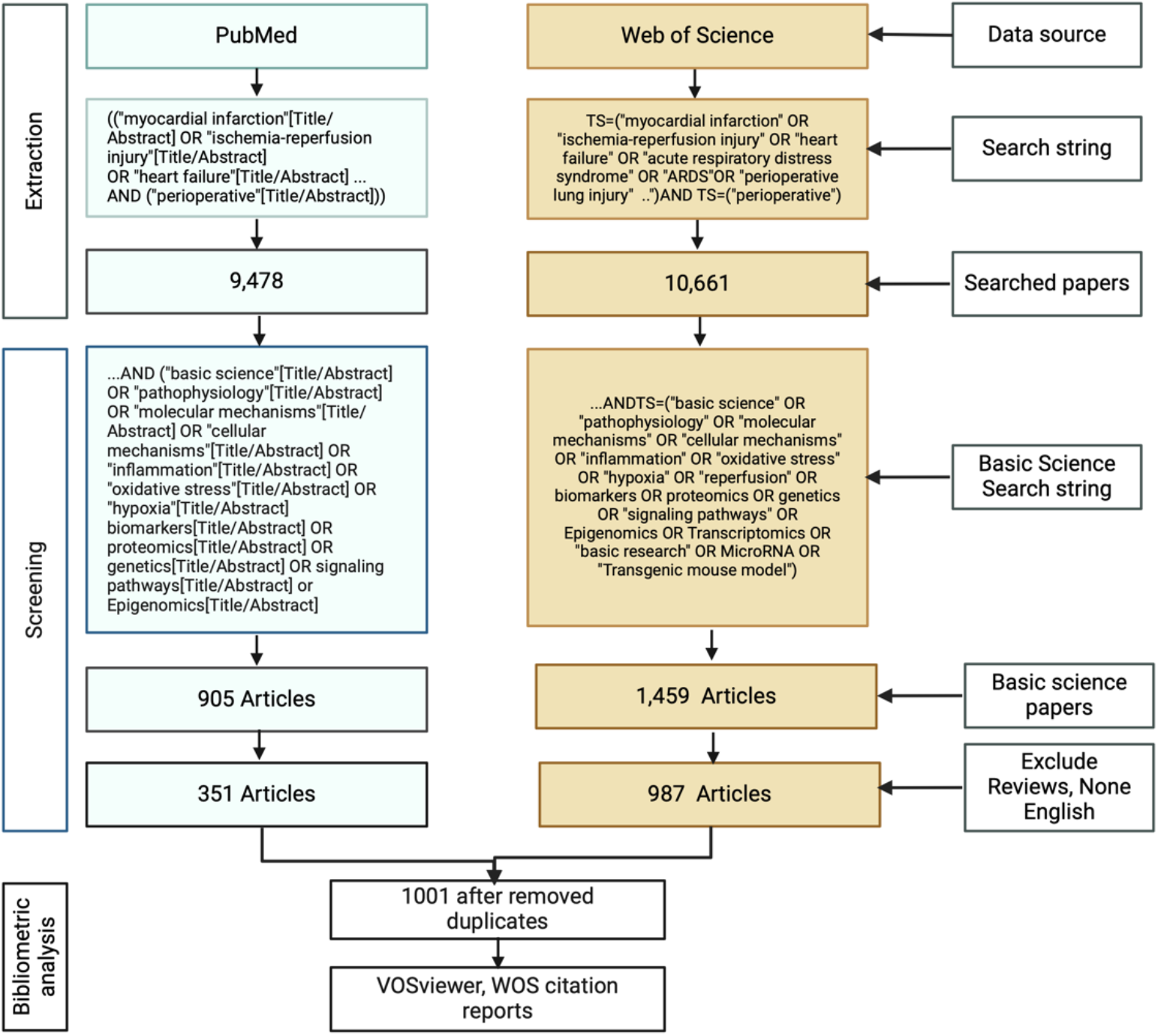
The workflow of the data extraction, screen and bibliometrics analysis.

The literature search was conducted using PubMed and Web of Science, chosen for their comprehensive coverage of scientific research and their robust search capabilities. To ensure the relevance and quality of the retrieved data, we limited the search to English-language publications, studies involving human subjects, and journal articles. The search results were then exported in the “Full Record and Cited References” format and saved as plain text files for further analysis.

Given that the term “perioperative organ injury” is too broad to effectively capture the basic science landscape in this field, clinical researchers advised focusing on more specific conditions commonly associated with perioperative organ injury. Therefore, our search strategy was tailored to include eight key conditions: “myocardial infarction,” “ischemia-reperfusion injury,” “heart failure,” “acute respiratory distress syndrome,” “perioperative lung injury,” “liver ischemia-reperfusion injury,” “ischemic acute kidney injury,” and “perioperative stroke.”

In PubMed, we employed MeSH (Medical Subject Headings) terms to refine the search, ensuring that the most relevant and specific articles were identified. MeSH terms allow for a more precise search by indexing articles according to standardized biomedical terminologies. For example, in searching for “ischemia-reperfusion injury,” we included MeSH terms related to cellular and molecular mechanisms, such as “Oxidative Stress,” “Inflammation,” and “Apoptosis,” to capture basic science research underlying this condition.

In Web of Science, we conducted an all-fields search using the same specific conditions as keywords. This approach ensured that we captured a broad spectrum of research, including articles that might not be indexed under MeSH in PubMed but are still relevant to the perioperative organ injury context. The search terms in Web of Science were designed to retrieve articles that covered both clinical and experimental research, with a focus on studies that explore the molecular pathways, gene expression patterns, and mechanistic insights related to the eight selected conditions.

By narrowing the search to these specific conditions, we aimed to obtain a more detailed and accurate overview of the basic science research landscape within the domain of perioperative organ injury. This approach allows us to identify the key areas of focus in the current literature and to understand the underlying mechanisms that contribute to organ injury in the perioperative setting.

### Inclusion and Exclusion Criteria

#### Inclusion Criteria

- Articles focused on the basic science mechanisms underlying perioperative organ injury.
- Studies discussing the pathophysiology, molecular, biomarkers, epidemiology, or cellular mechanisms of the specified conditions.
- Publications from peer-reviewed journals.
- Studies published in English.

#### Exclusion Criteria

- Reviews, editorial, case reports, commentaries and non-peer-reviewed content
- Studies not directly related to perioperative organ injury.
- Non-English publications.
- Data Extraction and Analysis.

We extract key studies that have significantly contributed to understanding the basic science of perioperative organ injury, data on publication trends, citation counts, research themes, keywords, countries, affiliations and author contributions from both PubMed and Web of Science. Merge papers after removed the duplicates from the two databases. We then used the collected data to perform bibliometric analysis, identifying key trends, research gaps, and influential papers in the field of perioperative organ injury basic science. We analyzed the publication trends over time to identify periods of significant research activities and emerging areas of interest. We did the co-occurrence analysis of keywords was conducted to determine the main research themes and potential gaps in the literature. Besides, we identified key contributors and influential papers in the field. Bibliometric analysis was performed using VOSviewer ^10^ which is designed for visualizing complex citation and co-authorship networks. and own scripts,

## Results

### 1. Publication trends of basic science of perioperative organ injury

The number of annual publications of perioperative organ injury and basic science research (1990-2024) was demonstrated in **Figure 2**. An increase in the number of publications over recent years indicates growing interest in the basic science of perioperative organ injury.

**Figure 2.**
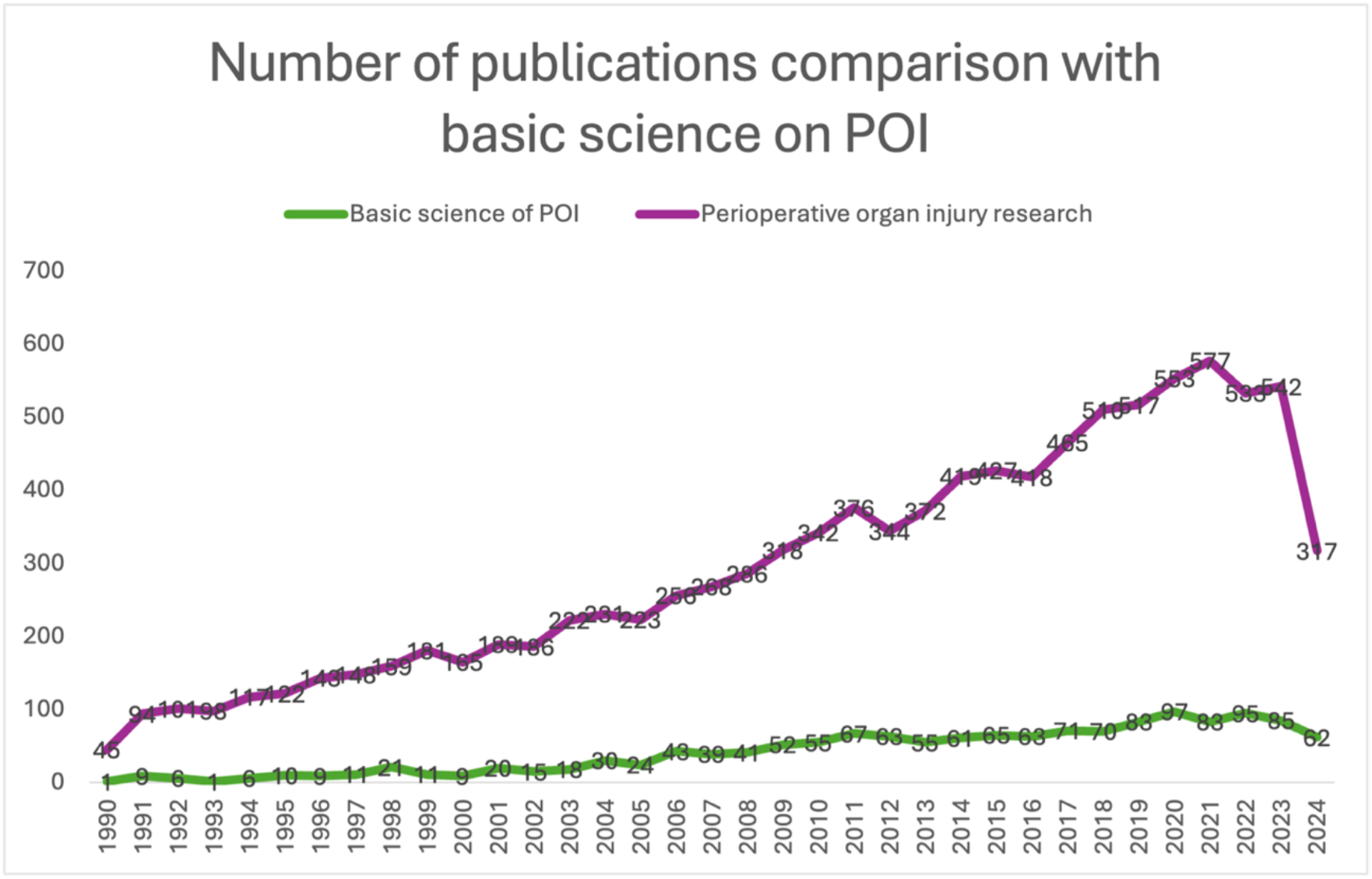
Comparison of Publications on Perioperative Organ Injury with Basic Science Since 1990. The trend shows a steady growth in basic science research within the field. The proportion of basic science studies in perioperative organ injury research has increased annually, rising from 2% in 1990 to an estimated 16% in 2024.

We checked the total publications per conditions, and found the research on these 8 conditions are imbalanced, certain area such as perioperative liver ischemia reperfusion injury remains underrepresented(see Table 1).

**Table 1.** Number of papers per search terms retrieved from WOS only.

| Number of papers | Search terms |
| --- | --- |
| 7,274 | perioperative(All Fields) AND stroke (All Fields) |
| 6,611 | perioperative (All Fields) and Myocardial infarction |
| 5,163 | perioperative (All Fields) and heart failure (All Fields) |
| 1,215 | perioperative (All Fields) and lung injury (All Fields) |
| 700 | perioperative (All Fields) and ischemia-reperfusion injury (All Fields) |
| 386 | perioperative(All Fields) AND ( acute respiratory distress syndrome (All Fields) OR ADRS (All Fields) ) |
| 299 | perioperative(All Fields) AND acute respiratory distress syndrome (All Fields) |
| 194 | perioperative (All Fields) and liver ischemia reperfusion injury (All Fields) |

To explore the relationships and intensity of research focus within the domain of perioperative organ injury, we analyze the co-occurrence of key terms extracted from literature data on perioperative organ injury and used VOSviewer to generate overlay, density and network visualization. Keywords, author suggested keywords and MeSH terms related to perioperative organ injury were extracted, with a minimum occurrence threshold of 10. A total of 563 keywords met this criterion. The analysis revealed six clusters of closely related terms in the overlay and network visualizations, indicating concentrated research areas. For example, terms related to pathway, expression, and myocardial ischemia reperfusion injury cluster together, highlighting a focused research area within the broader field of perioperative organ injury. **Figure 3** provides a comprehensive overview of the research landscape to identify key focus areas, trends in the literature on perioperative organ injury. Figure 3 A. is the research trends of perioperative organ injury basic science use the 8 specific conditions as examples to understand the changes of this field, the yellow nodes are the research focus in most recent years. For example, pathway, expression, mRNA, ROC, although they are smaller than green nodes, they are sparse and indicating the area are broadening, suggesting a growing interest in the molecular mechanisms of perioperative organ injury. Figure 3 B density visualization terms frequently appearing together in the literature are represented as dense clusters, with color intensity indicating the frequency and importance of these terms within the research corpus. Notably, areas related to injury, kidney injury and pathway appear as yellow clusters, indicating significant research activity, while terms associated with brain injury are less densely populated, suggesting potential gaps in the literature. This visualization not only highlights current research trends but also underscores the need for further investigation into less-explored aspects of perioperative organ injury.

**Figure 3.**
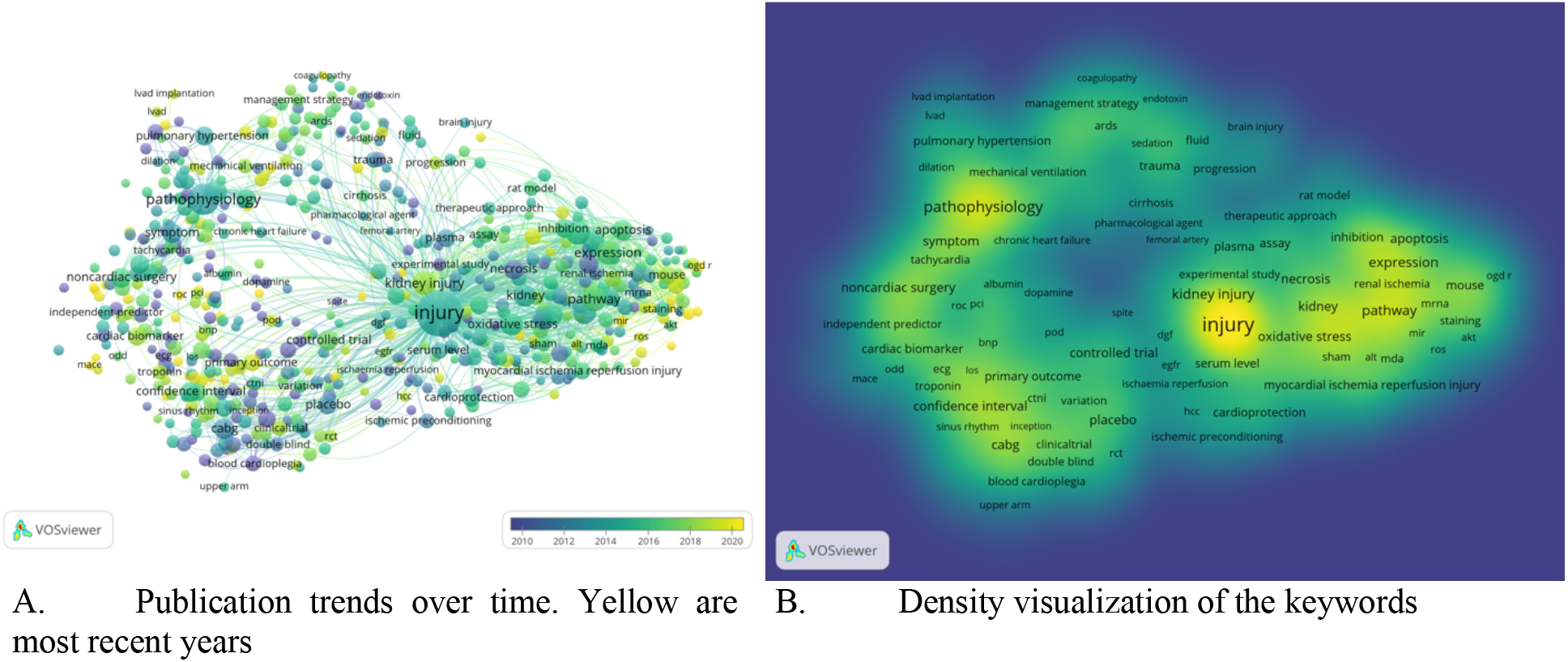
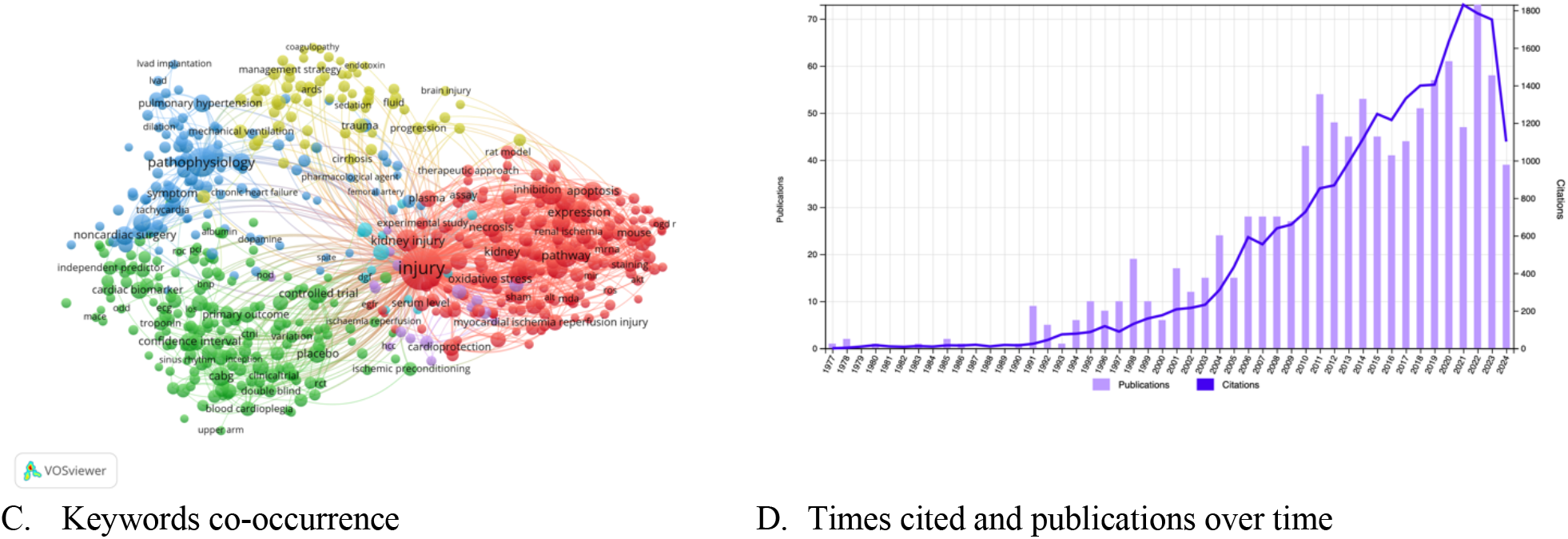
Bibliometric Visualizations of Perioperative Organ Injury (POI) Research. A. illustrates emerging research trends, with a growing focus on molecular mechanisms and pathways. B. presents a density plot highlighting key terms such as pathways, biomarkers, and predictors in POI research. C. shows a keywords co-occurrence map, identifying frequent research terms and their relationships. D. depicts the citation analysis of publications related to POI.

Figure 3C presents a network visualization generated by VOSviewer, illustrating the co-occurrence relationships between key terms. In this network, nodes represent specific terms, such as kidney injury, and pathway, while the links between nodes indicate the frequency and strength of co-occurrence in the literature. The size of each node reflects the frequency with which the term appears across the dataset, with larger nodes representing more commonly studied topics. Similarly, the thickness of the links between nodes indicates the strength of the relationship between terms—thicker links suggest a higher frequency of co-occurrence. Figure 3C also reveals distinct clusters grouped together based on their co-occurrence patterns. The colors of the clusters represent different thematic groups, providing an intuitive understanding of how research topics are organized within the field. The interconnected nature of these clusters underscores the complexity of perioperative organ injury and the multidisciplinary approaches required to advance knowledge in this field. Figure 3D illustrates the trends in the number of publications and their corresponding citation impact over time, based on data retrieved from the Web of Science database. The plot showcases the annual publication output alongside the total number of citations these publications have accrued each year, providing insights into both the growth of research activity and the influence of these studies in the field of perioperative organ injury. The number of publications has shown a steady increase since 1994, reflecting the growing interest and expansion of research in this domain. Notably, 2021 stand out with the peak of 2,805 citations, as shown in Figure 3D particularly in recent years more recent studies are not only more numerous but also highly impactful within the academic community. The overall trend of increasing citations and publications suggests a cumulative growth in knowledge, with newer studies building upon and expanding the findings of earlier research. This plot underscores the dynamic nature of perioperative organ injury research, where both the volume of research and its impact on the field are progressively increasing.

### 2. Geographical, affiliation and authors contribution of perioperative organ injury research

The bibliometric analysis of research contributions to the field of perioperative organ injury reveals a significant global engagement. To visualize the contributed countries and authors, we parsed the WoS RIS file to extract affiliations from the “AD” field, then remove duplicates and assign unique keys to each distinct affiliation. Convert these unique affiliations into geographic coordinates using a custom or in-house geocoding library. Integrate the geocoded data back into the RIS structure, associating each affiliation with its corresponding coordinates. Construct a coauthor network by creating edges between authors based on shared affiliations, and visualize the network on a geographic map, clustering the network based on geographic and coauthor ship relationships.

Figure 4. summarizes the importance of global participation in the research on perioperative organ injury. 66 countries were found has the publication of perioperative organ injury research. The Figure 4A diagram ranks the top contributing countries, highlighting the United States at the forefront, followed by China and the United Kingdom. These countries collectively account for a substantial portion of the research output in this field. Japan, Canada, South Korea, Turkey, and India also feature prominently, each contributing a significant number of publications. Australia rounds out the top contributors, reflecting the global interest and collaborative efforts in advancing knowledge and practices related to perioperative organ injury. This distribution underscores the importance of international collaboration and the diverse perspectives that drive innovation and improvements in this critical area of healthcare. Figure 4B highlights the top 40 institutions that have made significant contributions to the field of perioperative organ injury research. Harvard University emerges as a leading institution, with a substantial impact in advancing knowledge and practices in this area. The University of London and University College London also rank highly, reflecting their strong research programs and contributions to perioperative care. These institutions collectively drive innovation and clinical advancements, fostering a deeper understanding of perioperative complications and their management across the globe. Figure 4C depicts the co-authorship of POI research by clustering groups authors who frequently collaborate or publication together of closely related researchers. Figure 4D. Collaborative networks revealed strong connections between researchers in leading institutions, suggesting the importance of multidisciplinary collaboration in advancing the field. Collaborative networks revealed strong connections between researchers in leading institutions, suggesting the importance of multidisciplinary collaboration in advancing the field. This collaborative network diagram was generated by our in-house tool.

### 3. Leading journals and Citation Analysis

The most influential studies were those that provided novel insights into the mechanisms of perioperative organ injury and its implications for perioperative organ protection.

**Figure 4.**
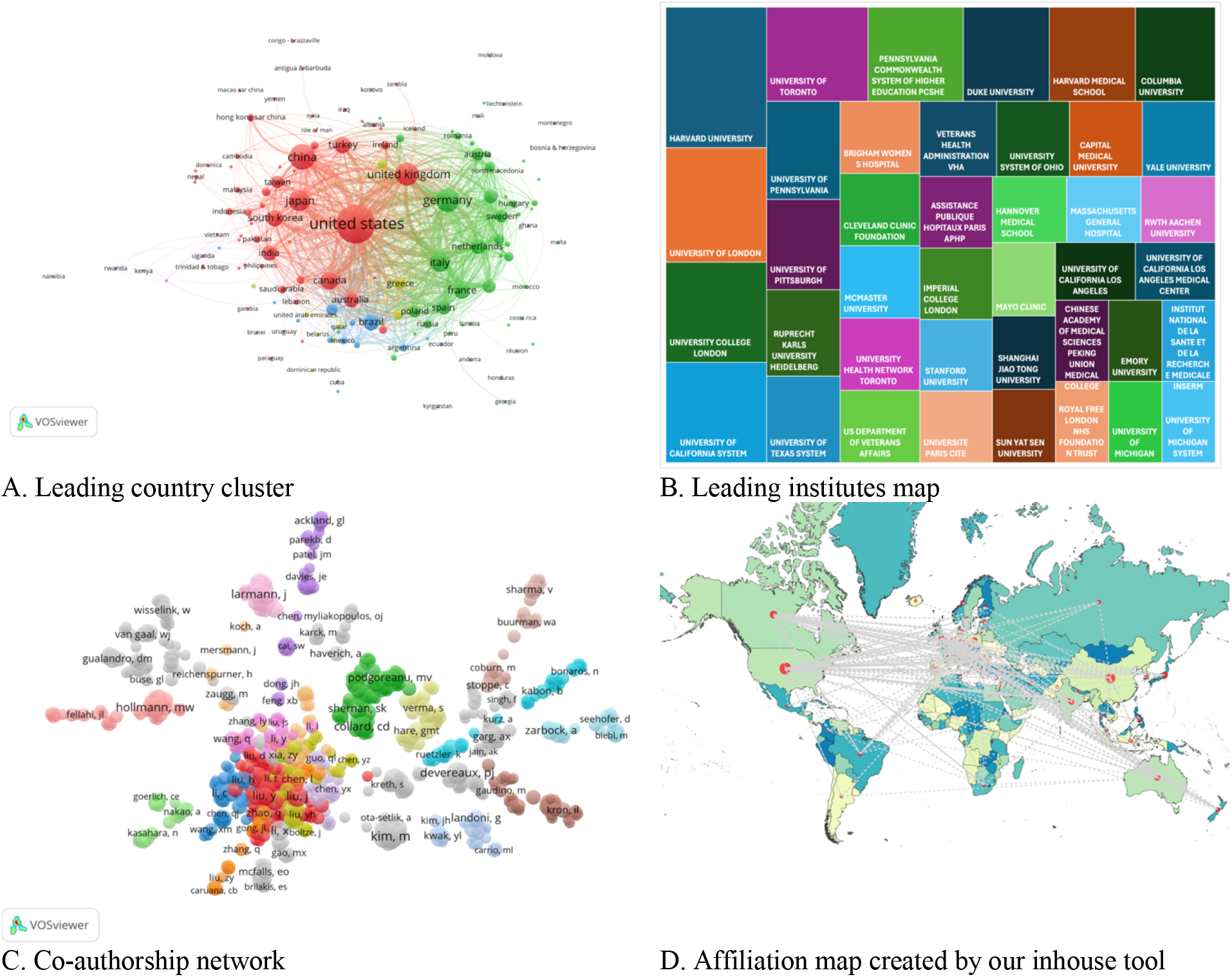
illustrates global contributors to perioperative organ injury research. A. presents a ranking of the top contributing countries, with the USA leading the list, followed by China, the UK, and Japan. B displays a map of the leading institutions in this field, highlighting Harvard University as the most prominent contributor. C shows a co-authorship diagram, illustrating the collaborative relationships between researchers. D features a geographic map of affiliations and countries, visualizing both the locations of institutions and their collaborative networks. In this map, nodes represent institutional affiliations, while edges indicate co-authorship connections between researchers.

The major journals with the most publications of perioperative organ injury and most citations were explored. Overall, 548 sorts of academic journals published articles and reviews about perioperative organ injury. As shown in Table 2, we observed that scientific achievements had a favored acceptance in these journals including journal of cardiothoracic and vascular anesthesia (n =43), Journal of thoracic and cardiovascular surgery (n =33) and Plos One(n=18). These top journals played a stronger role in this field.

**Table 2.** top 10 journals associated with POI.

| Rank | Journal Titles | Record Count |
| --- | --- | --- |
| 1 | Journal of cardiothoracic and vascular anesthesia | 43 |
| 2 | Journal of thoracic and cardiovascular surgery | 33 |
| 3 | Anesthesia and analgesia | 29 |
| 4 | Annals of thoracic surgery | 26 |
| 5 | Current opinion in anesthesiology | 24 |
| 6 | Journal of vascular surgery | 22 |
| 7 | European journal of cardio thoracic surgery | 21 |
| 8 | Anesthesiology | 20 |
| 9 | British journal of anesthesia | 19 |
| 10 | Plos One | 18 |

The analysis highlighted a small number of highly prolific authors and institutions that have significantly shaped the field. The most influential studies were those that provided novel insights into the mechanisms of ischemia-reperfusion injury and its implications for perioperative organ protection.

Table 3 summarized tope publication with more than citations in POI research. Key studies that have been widely cited were identified, underscoring their foundational role in the current understanding of perioperative organ injury.

**Table 3.**
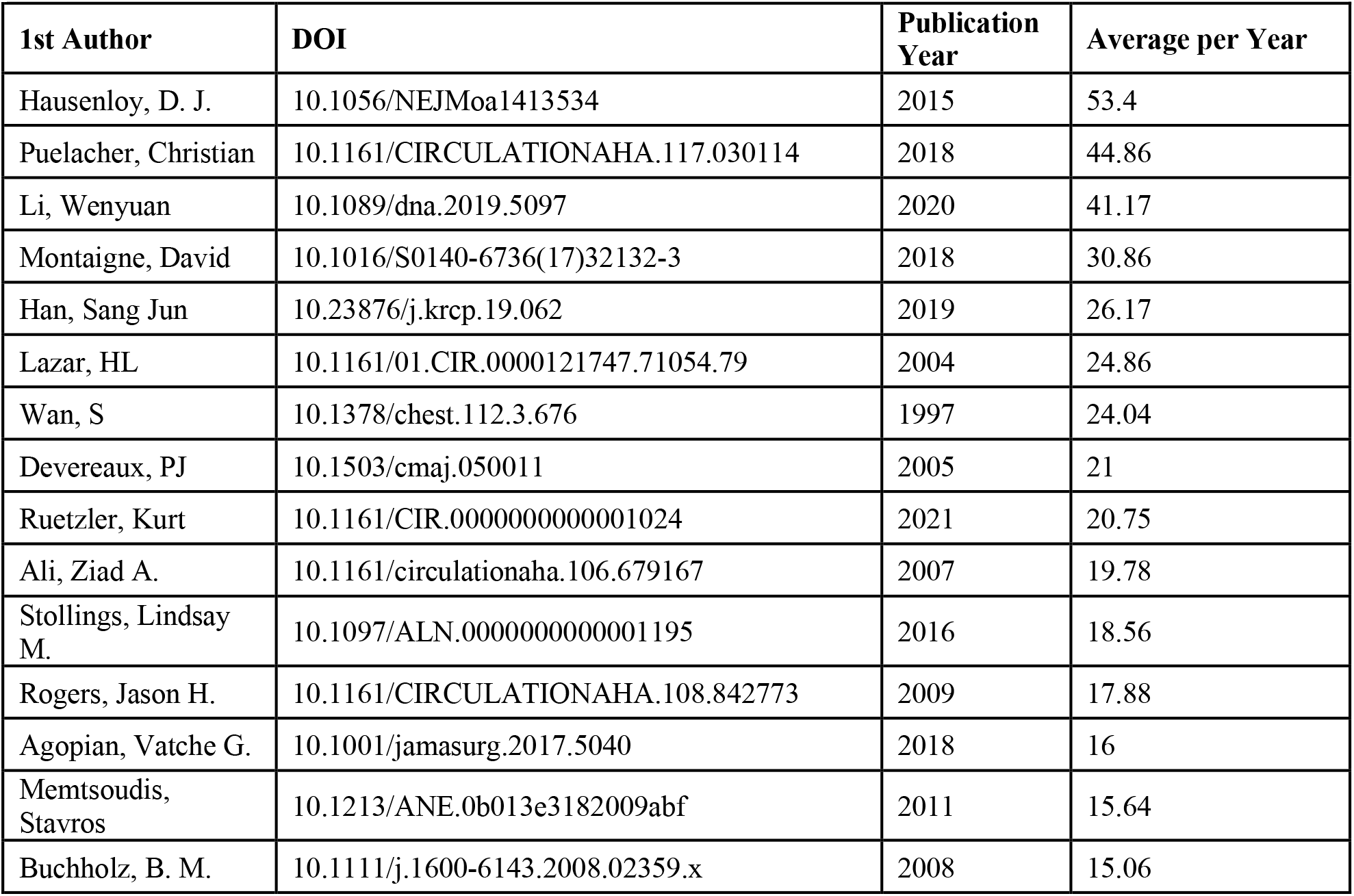
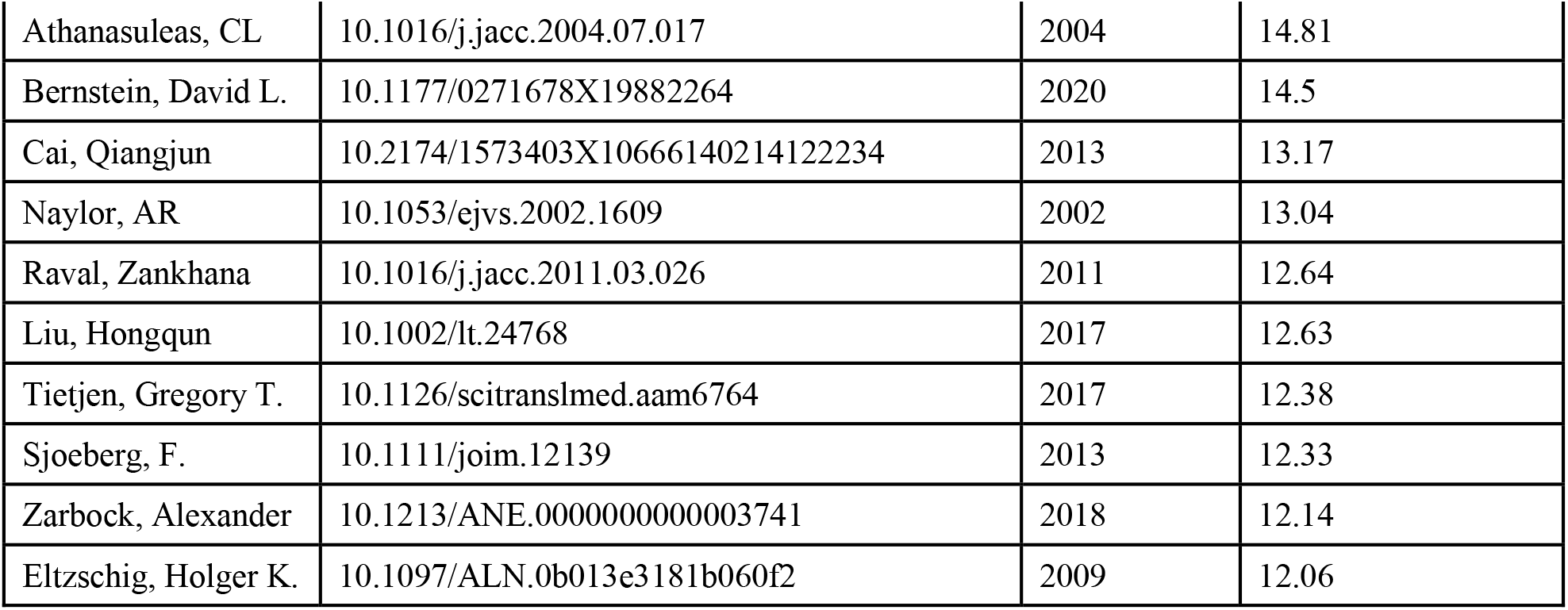
The top 25 publication in POI field with the first author, publication and the number of average citation per year.

## Discussion

The bibliometric analysis of perioperative organ injury research underscores the critical importance of basic science in advancing our understanding of this complex and multifaceted field. Over the past few decades, there has been a marked increase in research activity, particularly in exploring the underlying molecular mechanisms of perioperative organ injury. This shift is reflected in the clustering of keywords related to pathways, gene expression, and myocardial ischemia-reperfusion injury, underscoring the increasing emphasis on basic science research. These studies are crucial as they provide the foundational knowledge necessary to identify new therapeutic targets and develop more effective interventions. The transition from predominantly clinical reports to an emphasis on basic science reflects the maturation of the field and its recognition of the need to delve deeper into the biological processes that contribute to organ injury during surgery. However, the analysis also reveals that certain aspects of basic science research, such as the study of specific organ systems and the application of cutting-edge technologies like AI, machine learning, remain underexplored. Strengthening these areas is essential for a more comprehensive understanding of perioperative organ injury and for translating basic scientific discoveries into clinical innovations that can improve patient outcomes. As the field continues to evolve, it is imperative that future research prioritizes the integration of basic science with clinical practice, fostering interdisciplinary collaborations that can drive meaningful advancements in perioperative care.

The findings suggest that while significant progress has been made in understanding certain aspects of perioperative organ injury, there is still much to be done to translate these findings into effective clinical interventions. We acknowledges the limitations of bibliometric analysis, including the potential for bias due to the databases selected and the reliance on citation metrics as a proxy for research impact.

## Conclusion

This bibliometric analysis offers a comprehensive overview of the current research landscape on perioperative organ injury, with a particular emphasis on basic science. We are witnessing a significant surge in studies related to perioperative organ injury. The United States, China, and Europe have played pivotal roles in advancing this field. Notably, papers published in journals such as the Journal of Cardiothoracic and Vascular Anesthesia and the Journal of Thoracic and Cardiovascular Surgery have had a substantial impact. Our analysis shows a steady annual increase in research transitioning from clinical surgical reports to basic science investigations, including pathways, microRNA, and machine learning model. Strengthening basic science research in this domain is essential for deepening our understanding of perioperative organ injury and enhancing patient outcomes. Future research should prioritize the underrepresented areas identified in this study, with a focus on translating basic science discoveries into clinical practice.

## Data Availability

All data used in the paper are literate data available at PubMed, Web of Science

## Acknowledgement

This work was conducted under support from Cancer Prevention Research Institute of Texas (CPRIT) RR230020.

